# PERIPHERAL INOTROPES IN CRITICALLY ILL CHILDREN - IS IT SAFE?

**DOI:** 10.1101/2021.01.26.21250297

**Authors:** Ravi K Mooli, K Sadasivam

**Author notes:** **CORRESPONDING AUTHOR**: Dr K Sadasivam, Senior consultant, Paediatric Intensive Care Unit, Kanchi Kamakoti CHILDS Trust Hospital, 12A Nageswara Road, Nungambakkam, Chennai - 600 034.

## Abstract

Many children needing paediatric intensive care units care require inotropes, which are started peripherally prior to securing a central venous access. However, many hospitals in low- and middle-income countries may not have access to central lines and the vasoactive medications are frequently given through a peripheral venous access.

**Aim:** The aim of our study was to estimate the safety of peripheral vasoactive inotropes in children.

**Methods:** Children requiring peripheral vasoactive medications were included in this study. We retrospectively collected data at two time points on use and complications of peripheral vasoactive medications.

**Results:** Eighty-four children (51 pre-COVID era and 33 COVID pandemic) received peripheral vasoactive medications. Only 3% of children (3/84) developed extravasation injury, all of whom recovered completely.

**Conclusions:** Results from our study suggest that extravasation injury due to peripheral inotrope infusion is very low (3%) and it can be safely administered in children at a diluted concentration.

## INTRODUCTION

Many children needing paediatric intensive care units (PICU) care require inotropes, which are started peripherally prior to securing a central venous access. Although many PICU’s, health care centres and transport services have started using inotropes via peripheral intravenous catheters (1,2), the complications from peripherally administered vasoactive infusions in paediatrics are not very well described (3). In addition, many health care centres in low- and middle-income countries (LMIC) do not have facilities or access for using central venous catheters. Here we describe a study assessing the safety of peripheral vasoactive inotropes at a large PICU in Chennai, India.

## METHODS

We retrospectively collected data from 1^st^ January 2019 to 1^st^ October 2019 and again from 1^st^ April 2020 to 1^st^ October 2020 (during the COVID 19 pandemic). All children who received vasoactive mediations through peripheral intravenous catheter (PIC) were included. We described complication as tissue injury, which included any erythema, blistering, skin breakdown, or necrosis. Peripheral inotrope infusion concentration used was lower than the concentration of central inotrope infusion (Table 1). The concentration was decided and agreed by the PICU pharmacist and PICU consultants. Ethical approval for the study was obtained from the hospital ethical committee.

**Table 1:**

| **Characteristics of children requiring vasoactive medications via PIC** |  |  |
| --- | --- | --- |
|  | **Pre-COVID 19 era**<br>**n=51** | **COVID 19 era**<br>**n=33** |
| **Age in years (range)** | **2.6 (1 month – 18 years)** | **7 (IQR, 5 – 7)** |
| **Male (n)** | **25** | **16** |
| **Female (n)** | **26** | **17** |
| **Admitted from (n)** |  |  |
| *Emergency room* | 37 | 23 |
| *Wards* | 9 | 6 |
| *Other hospital* | 5 | 4 |
| **Indication for vasoactive medications (n)** |  |  |
| *Septic shock* | 20 | 3 |
| *Dengue shock* | 17 | 0 |
| *Cardiogenic shock* | 10 | 2 |
| *Hypovolemic shock* | 1 | 0 |
| *Traumatic brain injury* | 3 | 1 |
| *Cardiogenic shock (PIMS-TS\* related)* | 0 | 27 |
| **Length of PICU stay in days (IQR)** | **3 (0.25 - 27)** | **3 (IQR, 2 – 5)** |
| **Type of vasoactive medication (n) %** |  |  |
| *Epinephrine (0.15mg/kg in 50 ml Saline)* | (35) 67% | 6 (18 %) |
| *Nor-epinephrine (0.15mg/kg in 50 ml Dextrose)* | (33) 65% | 27 (81%) |
| *Dobutamine (6mg/kg in 50 ml Saline)* | (6) 12% | 0 (0%) |
| *Milrinone (1.5mg/kg in 50 ml Dextrose)* | (4) 8% | 2 (6%) |
| *Dopamine (6mg/kg in 50 ml Saline)* | (4) 8% | 0 (0%) |
| **Mean duration of vasoactive mediations via PIC (hr, range)** | **12 (1 -72)** | **8 (1 – 48)** |
| **Complications (n) %** |  |  |
| *Extravasation injury* | (3) 6% | 0 |
| **Children needing central venous access n (%)** | **21 (41%)** | **2 (6%)** |

\*PIMS-TS: Paediatric inflammatory multisystem syndrome temporarily associated with SARS-COV2
PIC: peripheral intravenous catheters

## RESULTS

Fifty-one children from pre-COVID 19 period and 33 children during the COVID 19 pandemic were included. Demographic and clinical characteristics of all children are shown in Table 1. In all 84 children, 22-gauge peripheral venous catheter was used and only three children developed extravasation injury, all of which were local tissue necrosis. They did not require any intervention and recovered completely.

## DISCUSSION

Central venous access (CVA) is commonly preferred over PIC for giving vasoactive medications (1,2,4). However, in an emergency, CVA in children can be challenging and difficult, delaying the administration of vasoactive medications (5). Using a PIC may be beneficial in such situations and lifesaving (2,5).

Adult published studies have documented extravasation injury tends to occur due to long duration of infusion via PIC (2,5). Our findings appear to be in consistent with this. In all three children who developed extravasation injury, vasoactive mediations were given for more than 24 hours. Their average PICU stay was 17 days and median infusion time was 28 hours, indicating that duration of infusion may be a risk factor for PIC injuries.

While administering vasoactive mediations through PIC, close and frequent site monitoring is necessary for timely detection of extravasation injury (1,2,4). In our PICU, children receiving vasoactive mediations via PIC are monitored hourly for any signs of tissue injury using standard phlebitis and infiltration scale. There are no available studies to suggest if any particular vasoactive medication or the concentration of infusion increases the risk of injury. Although extravasation injury due to peripheral inotrope infusion in our study is very low (3%) we recommend using a diluted concentration of vasoactive medications along with frequent assessment of the limb to identify any risks, hence minimising the complications.

In children requiring shorter duration of vasoactive medications, using a PIC may be cost effective, especially in LMIC, in addition to minimising the complications of CVA. During the current COVID 19 pandemic we have successfully administered vasoactive medications through PIC in children presenting with Paediatric inflammatory multisystem syndrome temporally associated with COVID-19 (PIMS-TS), thus avoiding the need for general anaesthesia and CVA.

Our study adds to the available evidence that vasoactive medications at a diluted concentration can be safely administered through PIC in children who require short duration of vasoactive medications. This study is beneficial for clinicians in LMIC and adds to exiting evidence that peripheral inotrope usage is safe with adequate monitoring.

## Data Availability

The datasets generated during and/or analysed during the current study are available from the corresponding author on reasonable request

## ACKNOWLEDGEMENTS

No specific funding was received for this study.

